# Comprehensive Genetic Profiling of Sensorineural Hearing Loss Using an Integrative Diagnostic Approach

**DOI:** 10.1101/2024.12.08.24318682

**Authors:** Sang-Yeon Lee, Seungbok Lee, Seongyeol Park, Sung Ho Jung, Yejin Yun, Won Hoon Choi, Ju Hyuen Cha, Hongseok Yun, Sangmoon Lee, Myung-Whan Suh, Moo Kyun Park, Jae-Jin Song, Byung Yoon Choi, Jun Ho Lee, Young Seok Ju, June-Young Koh, Jong-Hee Chae

## Abstract

Despite the advent of Next-Generation Sequencing (NGS), genetic diagnosis of genetic disorders remains challenging, with diagnostic rates plateauing at approximately 50%. We investigated sensorineural hearing loss (SNHL), a prevalent sensory disorder with substantial genetic heterogeneity, through a comprehensive genomic analysis of a homogeneous disease cohort. Leveraging 394 families (750 individuals), we implemented a systematic multi-tiered genomic approach encompassing single-gene analysis to whole-genome sequencing (WGS), integrated with functional assays and bioinformatic analysis. Our methodological framework revealed a cumulative diagnostic yield of 55.6% (219 families), with automated WGS bioinformatics pipeline uncovering an additional 20 families harboring pathogenic variants, predominantly structural variants. Notably, comparative genomic analysis unveiled a higher frequency of single pathogenic alleles in recessive genes within our SNHL cohort relative to control populations. Subsequent deep intronic region interrogation identified three pathogenic variants on the opposite allele, substantiating the diagnostic utility of comprehensive genomic profiling. Through this approach, we delineated a genome-phenome landscape of SNHL, elucidating molecular signatures and establishing genotype-phenotype correlations at the inner ear functional level. This study underscores the transformative potential of WGS as a robust molecular diagnostic modality, advancing precision medicine paradigms in genetic disease research.

## Introduction

Hearing is the primary sense used for human communication and an important component in the development of language and music. Thus, hearing impairment, the most common sensory deficit in humans, is a major public health problem, affecting approximately 466 million people worldwide (World Health Organization, https://who.int/news-room/fact-sheets/detail/deafness-and-hearing-loss). Sensorineural hearing loss (SNHL)—i.e., defective sound signaling in the auditory sensory system—can be caused by multiple etiologies, including genetic causes, congenital infections, trauma, ototoxic medications, and autoimmune disorders^1^. Since the 2010s, advances in high-throughput next-generation sequencing (NGS) technologies have facilitated extensive elucidation of the genetic backgrounds of SNHL, with a focus on monogenic forms of deafness. Notably, mouse genetics studies have helped reveal the physiological basis of SNHL in humans and the associated molecular functions^2^.

Despite growing recognition of the significance of genetic diagnosis of SNHL, it remains challenging to identify a genetic diagnosis in SNHL with substantial genetic heterogeneity^3,4^. NGS is increasingly favored for genetic diagnosis due to its capacity for simultaneous large-scale genetic loci screening, and methods like targeted panel sequencing (TPS) and whole-exome sequencing (WES) were widely used in real-world practice^5–7^. In the literature, targeted sequencing for SNHL has achieved diagnostic yields of between 12.7% and 64.3%^8–11^. However, even after exome sequencing, approximately 50% of cases remain genetically elusive.

As the cost of sequencing dramatically declines^12^, the clinical application of whole-genome sequencing (WGS) becomes more feasible, which has a higher capability to detect a more diverse spectrum of genomic variants that had not previously been captured by exome sequencing or other targeted approches^13,14^. Recent studies have shown the clinical utility of WGS for the genetic diagnosis of several disorders and effectively shortening their diagnostic odyssey, increasingly considered as a first-line genetic test^15–19^. However, WGS is not yet widely applied in routine clinical settings for diagnosing patients with rare diseases, including SNHL, due to several limitations, such as the difficulties of rapid bioinformatic analysis and accurate clinical interpretation. Additionally, although whole genomes are sequenced, the analysis is often limited to in silico gene panels or the coding regions of the genome^20^.

In the present study, we comprehensively explored the genetic landscape of 394 prospective SNHL families. Using a stepwise approach from single target gene analysis to WGS, we evaluated the additional diagnostic value of WGS. We implemented an automated WGS bioinformatics pipeline, integrating *in-house* algorithms with manual curation by both otologists and medical geneticists. This approach allowed comprehensive analysis of all variant types, and improved the diagnostic yield for previously undiagnosed patients. Further analysis of deep intronic regions identified novel pathogenic variants. These findings refined the genotype-phenotype landscape of SNHL, revealing the gene signatures based on phenotypes and identifying genotype-phenotype correlations at the level of inner ear molecular functions. Our results demonstrate the clinical utility of an integrated molecular diagnostic approach, including WGS, in real-world SNHL practice, paving the way toward precision medicine.

## Results

### A stepwise approach of genetic testing for patients with SNHL

We conducted stepwise approach of genetic testing, sanger sequencing, TPS, WES, mitochondrial DNA (mtDNA) sequencing, multiplex ligation-dependent probe amplification (MLPA), and WGS, to prospectively recruited SNHL cohort (*n* of probands = 394; n of participants including probands and their family members = 750; **Supplementary Table 1**), including non-syndrome hearing loss (ns-SNHL; *n* = 341, 86.5%), and syndromic hearing loss (s-SNHL; n = 53, 13.5%). Genetic testing was structured into five sequential steps (**Fig. 1 and Supplementary** Fig. 1a). In Step 1, we screened 22 variants from 10 classical deafness genes (*GJB2, SLC26A4*, *TMPRSS3*, *CDH23*, *OTOF*, *TMC1*, *ATP1A3*, *MPZL2*, *COCH*, and 12S rRNA). Among cohort, ns-SNHL patients (*n* = 341) were initially subjected to Step 1 and the ns- SNHL patients who remained undiagnosed after Step 1 (*n* = 295), and s-SNHL patients (*n* = 53) were then subjected to the next steps (Steps 2-1 and 2-2). In Step 2-1, 99 and 249 patients with SNHL were subjected to TPS (including 246 genes) and WES, respectively. After that, undiagnosed patients with suggestive clinical features, including symmetric mild-to-moderate SNHL (*n* = 51)^21^, apparent branchio-oto-renal/branchio-otic (BOR/BO) syndrome (*n* = 2)^22^, radiological evidence of enlarged vestibular aqueduct (EVA) (*n* = 1), and suspicious mitochondria disorders (*n* = 11) were subjected to Step 2-2, including MLPA methods (*n* = 54) or mtDNA panel sequencing (*n* = 11). In Step 3-1, among patients who remained undiagnosed, all s-SNHL patients and a representative subset of ns-SNHL patients were selected for WGS using a well-thought-out sample size estimation with a stratified sampling approach (**Supplementary** Fig. 1b). For the remaining undiagnosed patients (*n* = 100), deep intronic regions of SNHL-related genes were screened to identify additional pathogenic variants in Step 3-2. After stepwise genetic testing, patients with suspected pathogenic genetic causes underwent subsequent bioinformatic analyses and curation. Multidisciplinary molecular board meetings, comprising both clinicians and genome scientists, were then conducted to confirm genetic diagnosis.

**Figure 1.**
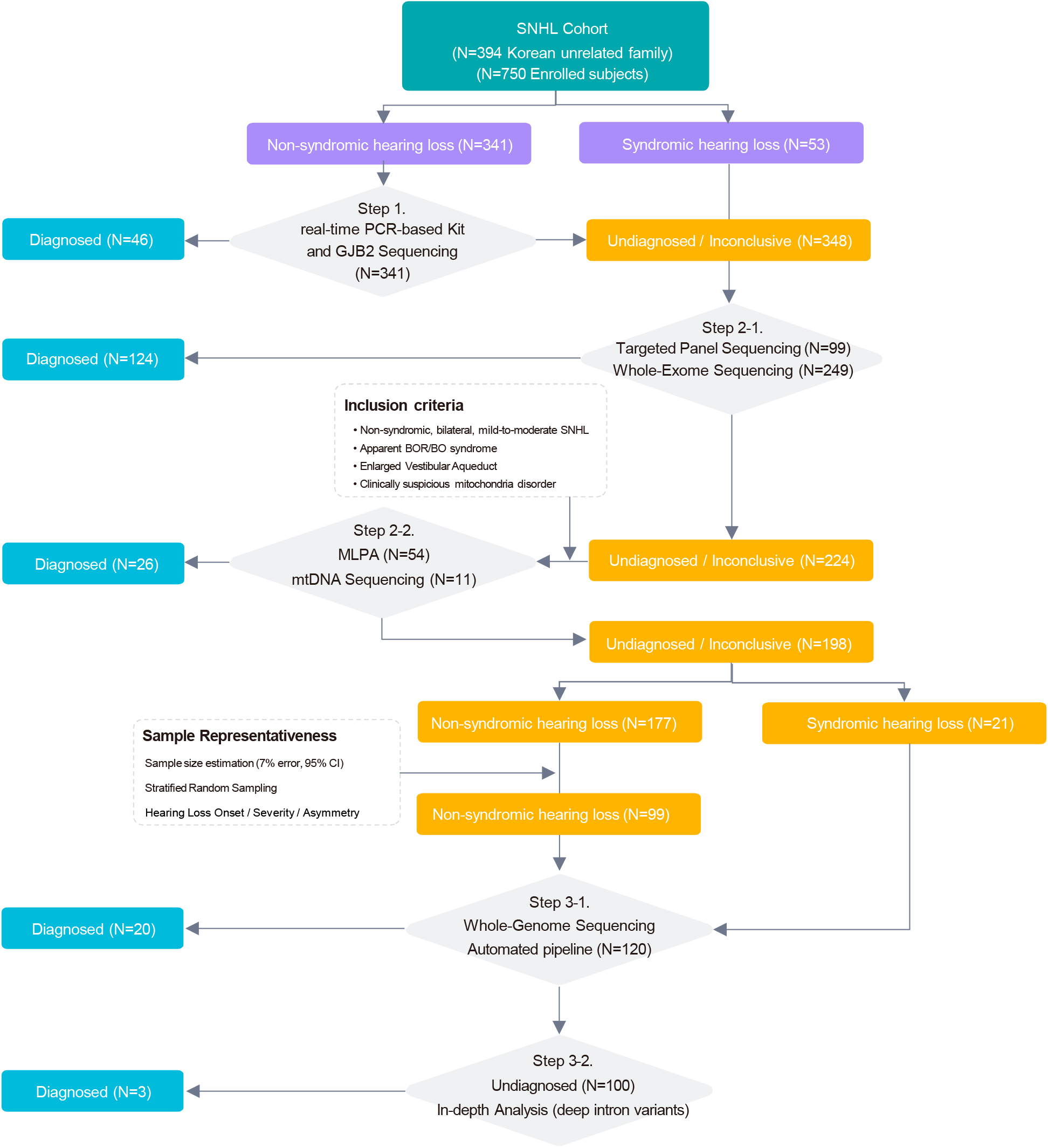
Study design and diagnostic pipeline. Flow diagram illustrating a prospective, step-by-step genetic approachof 394 unrelated SNHL families and 750 individuals including probands in our cohort study. The diagnostic pipeline included Step 1 (real-time PCR screening kit and direct *GJB2* sequencing), Step 2-1 (targeted panel sequencing or whole-exome sequencing), Step 2-2 (MLPA and/or mtDNA panel sequencing), Step 3-1 (whole-genome sequencing), and Step 3-2 (deep intronic variant analysis). SNHL, sensorineural hearing loss; BOR/BO syndrome, branchio-oto-renal/branchio-otic syndrome; MLPA, multiplex ligation-dependent probe amplification; mtDNA, mitochondrial DNA; CI, confidence interval.

### Incremental improvement in diagnostic yield through stepwise genetic testing for SNH*L*

Through comprehensive genetic testing, we incrementally improved the diagnostic yield in the cohort, identifying disease-causing variants in 219 families (55.6%; **Fig. 2a**). Following the stepwise approach, in Step 1, we identified causal variants in 46 out of 341 patients (diagnostic yield for Step 1: 13,5%; cumulative yield: 11.7%). In Step 2-1, causal variants were found in 124 out of 348 patients (diagnostic yield for Step 2-1: 35.6%; cumulative yield: 43.1%), while Step 2-2 revealed causal variants in 26 out of 65 patients (diagnostic yield for Step 2-2: 40%; cumulative yield: 49.7%). WGS was performed on 120 patients, identifying causal variants in additional 23 probands (20 in Step 3-1 and 3 in Step 3-2; diagnostic yield for Step 3: 19.2%; cumulative yield: 55.6%). Details of individual patients, including the performed tests, diagnostic outcomes, and identified variants, are summarized in **Supplementary Table 2**.

**Figure 2.**
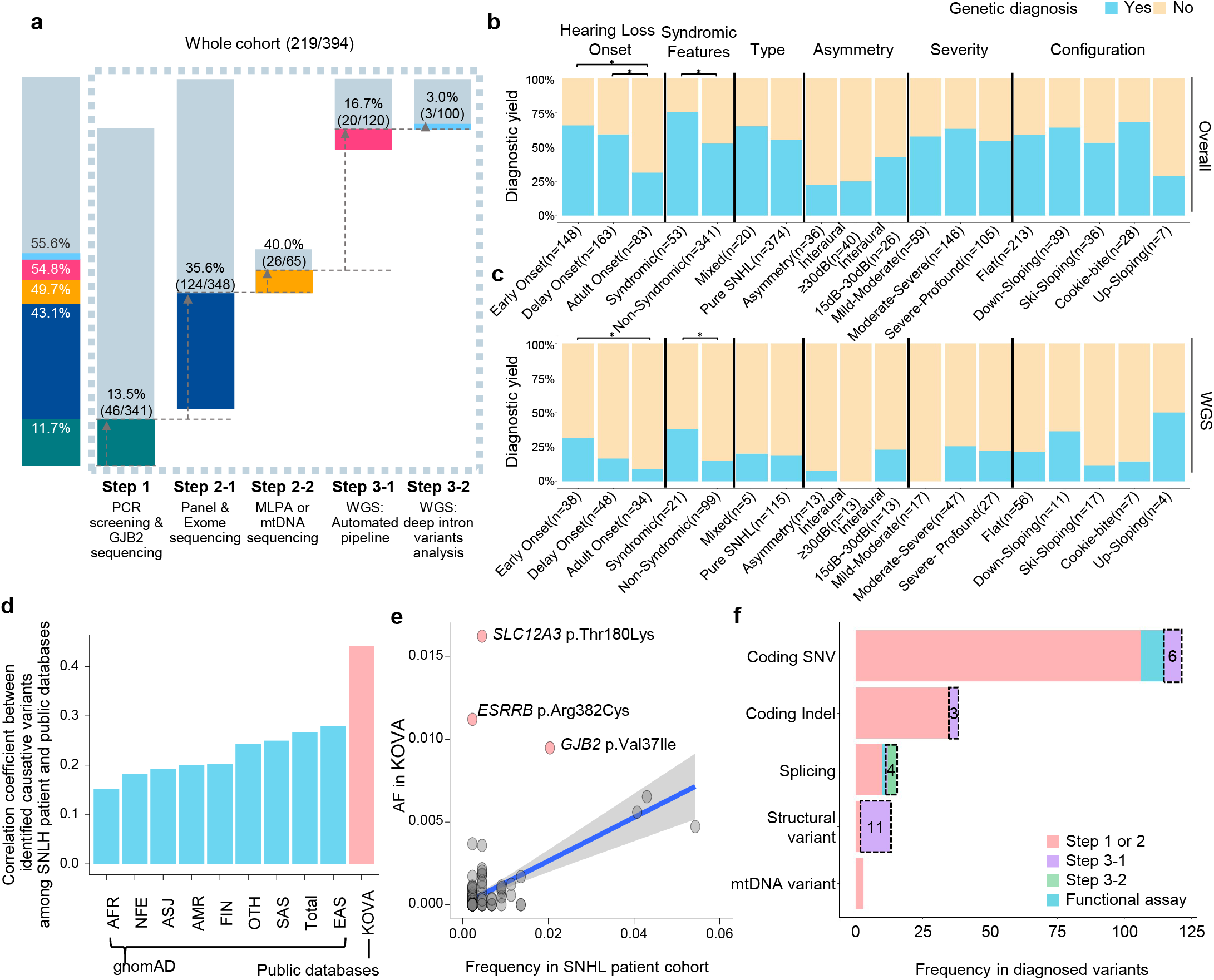
Stepwise genetic diagnosis outcomes in SNHL patients. (a) Diagnostic yield of each genetic test for the whole SNHL cohort. Bar graph showing the cumulative diagnostic rate according to genetic diagnosis steps. (b-c) Diagnostic yield based on SNHL phenotypes and comparative analysis within the whole SNHL cohort (*n* = 394 families; b) and within the WGS cases (*n* = 120 families; c). Statistical significance for hearing loss onset and syndromic features was determined using one-way ANOVA with Bonferroni’s multiple comparisons tests and the t-test, respectively. Significance levels are indicated as \**P* < 0.05. (d) Pearson’s correlation coefficient values for causative variants between the allele frequencies (AFs) in our cohort and those from other populations. The AFs in our cohort showed the highest correlation with the Korean database, KOVA, followed by the East Asian population within gnomAD. (e) Dot plots shows the AF correlation between our cohort and KOVA. There are three variants (pink dots) showing higher AFs in KOVA, suggesting their phenotypic variability such as low penetrance or late onset associated with these variants. A fitted line of linear regression model (blue line) and 95% confidence intervals (grey area) are displayed. (f) Distribution of variant subtypes identified at each diagnostic step. WGS, whole-genome sequencing; MLPA, multiplex ligation-dependent probe amplification; mtDNA, mitochondrial DNA; SNHL, sensorineural hearing loss; SNV, single nucleotide variant; indel, insertion/deletion.

Multivariate analysis revealed that the rate of genetic diagnosis was higher among patients with early identification of SNHL (adjusted odds ratio [OR], 1.32; 95% confidence interval [CI], 1.11–1.57) and those with a family history (adjusted OR, 1.55; 95% CI, 1.31–1.82; **Fig. 2b and Supplementary Table 3**). Similarly, patients with syndromic features were more likely to have identified genetic variants (adjusted OR, 1.44; 95% CI, 1.17–1.70). In contrast, a lower rate of genetic diagnosis was observed in patients with adult-onset SNHL (adjusted OR, 0.50; 95% CI, 0.35–0.68), asymmetric hearing loss (adjusted OR, 0.38; 95% CI, 0.18–0.64), and interaural asymmetry (adjusted OR, 0.43; 95% CI, 0.23–0.68). Additionally, in the context of WGS, several features were associated with a higher likelihood of achieving a genetic diagnosis (**Fig. 2c and Supplementary Table 4**). The multivariate analyses suggested that genetic diagnosis was significantly related to the presence of syndromic features (adjusted OR, 2.51; 95% CI, 1.15–5.04) and early identification through a failed newborn hearing screening (NHS) (adjusted OR, 2.35; 95% CI, 1.13–4.97). In addition, the diagnostic yield significantly varied depending on the WGS approach. Trio-based WGS had a higher likelihood of identifying causal variants (adjusted OR, 3.71; 95% CI, 1.67–9.68), whereas singleton WGS was less effective (adjusted OR, 0.32; 95% CI, 0.12–0.71).

Further, we conducted the correlation analysis between the frequencies of causative variants among our cohort and population allele frequencies of the variants from public databases, such as the gnomAD^23^ and KOVA^24^ (Korean Variant Archive: Korean population database; **Fig. 2d**). We observed the strongest association with East Asians compared to other ethnicity groups within gnomAD, and a notably higher association with KOVA, suggesting that patient ethnicity should be considered in variant discovery. We discovered three outlier variants (*SLC12A3* p.Thr180Lys, *ESRRB* p.Arg382Cys, and *GJB2* p.Val37Ile) when comparing the variant frequencies with allele frequencies in KOVA (**Fig. 2e**). These variants have higher frequencies in the Korean population than in diagnosed patients, suggesting either their low penetrance features or the potential for functional pathogenicity despite its particularly higher allele frequencies^25^.

In conclusion, from Step 1 to Step 3-2, we identified the genetic causes of SNHL in 219 out of 394 probands (55.6%), with 23 diagnoses made through WGS. Among the identified variants, deep intronic variants (*n* = 3) and structural variants (SVs) (*n* = 11) were predominantly detected through WGS, and 13 variants of uncertain significance (VUS) required functional assays to validate their pathogenicity (**Fig. 2f**).

### Comprehensive characterization of causative variants in SNHL

Using the genetic findings obtained through comprehensive stepwise genetic tests, we illustrated a mutational landscape of SNHL. Collectively, 63 genes were identified as disease-causing within 219 genetically diagnosed families (**Fig. 3a and Supplementary** Fig. 2a). *GJB2* was the most frequently affected gene (10.5%, 23/219), followed by five genes (*SLC26A4*, *STRC*, *USH2A*, *CDH23*, *and MPZL2)* that were found in at least ten unrelated families (collectively >40% of all diagnosed cases). Conversely, 29 SNHL-associated genes were detected from only one family (collectively 13.2%; 29/219; **Supplementary** Fig. 2a), suggesting that many more rare genes can cause SNHL. The inheritance patterns of the 63 genes included autosomal recessive (26/63 genes; affecting 132 families; found double hits, including homozygote and compound heterozygote variants), autosomal dominant (33/63 genes; affecting 73 families; found single hit), X-linked (4/63 genes; affecting 6 families), and mitochondrial (3/63 genes; affecting 8 families) (**Fig. 3a**). Within our cohort, three identified genes—*TECTA*, *COL4A3*, and *WFS1*—exhibited both autosomal recessive and autosomal dominant inheritance traits. Moreover, one patient exhibited dual genetic etiologies, inherited from their parents, harboring compound heterozygous variants (c.299del:p.His100LeufsTer12 and c.235del:p.Leu79CysfsTer3) in *GJB2* and a heterozygous variant (c.113G>A;p.Gly38Asp) in *COCH*.

**Figure 3.**
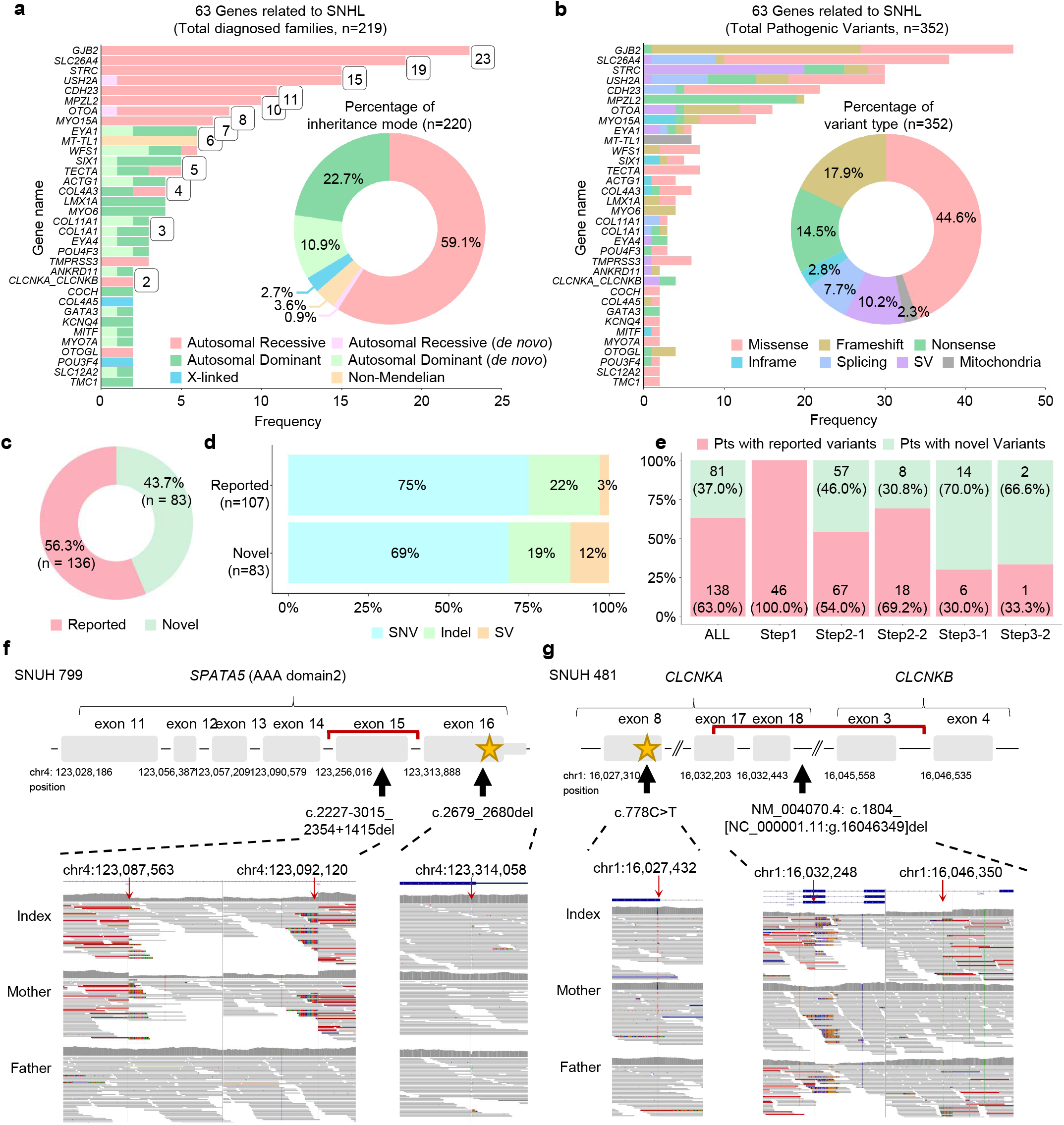
Genomic landscape of the SNHL cohort. (a) Bar plot showing the frequencies and inheritance patterns of 63 SNHL-associated genes from 219 genetically diagnosed families. Pie chart showing the percentages of inheritance patterns. (b) Bar plot showing the mutational landscape of the total 352 likely pathogenic or pathogenic variants among the 63 SNHL genes. Pie chart showing the percentages of variant types. (c) Proportion of novel variants among identified causal variants. (d) Structural variants (SVs) were more common among novel variants compared to previously reported variants. (e) Novel variants were frequently identified through WGS (Step 3-1) and SpliceAI-based deep intronic variants analysis (Step 3-2). (f, g) Schematic illustrations showing the location of each identified pathogenic variant within *SPATA5* and *CLCNKA-CLCNKB* in the probands (top, respectively). The genomic regions corresponding to each variant are visualized using the integrative genomics viewer (bottom) for each figure. SNHL, sensorineural hearing loss; SNV, single nucleotide variant; indel, insertion/deletion; SV, structural variation.

**Fig. 3b** and **Supplementary** Fig. 2b display the mutational landscapes of the 352 causative variants, including the variant type. Among the 352 identified variants, we found missense variants (157, 44.6%), nonsense variants (51, 14.5%), frameshift variants (63, 17.9%), and inframe variants (10, 2.8%) within the exome region. Additionally, splicing variants (27, 7.7%), SVs (36, 10.2%), and mitochondrial variants (8, 2.3%) were identified. Interestingly, we observed differences in the distribution of variant types across genes. In particular, consistent with previous reports, SVs accounted for (20/30, 66.7%) of the total variants in the *STRC* gene.

Among the 352 causative variants (190 when the same variants were collapsed), 83 variants were novel (**Fig. 3c**). All of the 352 variants were either pathogenic (P) or likely pathogenic (LP) according to the American College of Medical Genetics and Genomics (ACMG) and Association for Molecular Pathology (AMP) guidelines (**Supplementary Table 5**)^26^. A variety of functional studies— ranging from molecular modeling to minigene splicing assay—were performed to test 16 highly suggestive variants (**Supplementary Table 6**), leading to the reclassification of 13 variants from “uncertain significance” to “likely pathogenic”. An analysis of the variant type distribution among the 83 novel variants revealed that SVs, detected mostly through WGS, accounted for a much larger proportion (12.0%) compared to previously reported variants (2.8%) **(Fig. 3d)**. In addition, we observed an increasing trend in the proportion of patients with novel variants as the steps progressed from Step 1 to Step 3-2 (**Fig. 3e**).

Novel SVs identified from WGS include exonic deletions on *SPATA5* and *CLCNKA-CLCNKB* (**Figs. 3f and 3g**). In one patient (SNUH 799), WGS identified a small SV (c.2227-3015_2354+1415del) exclusively involving exon 15 of *SPATA5* (**Fig. 3f**), along with an *in trans* short frameshift deletion in exon 16 (c.2679_2680del). Notably, consistent with a previous case report^27^, this patient exhibited systemic clinical manifestations, including bilateral moderately severe SNHL, intractable epilepsy with diffuse brain atrophy, and global developmental delay (**Supplementary** Fig. 3a). Next, we investigated the molecular consequences of the small SV (c.2227- 3015_2354+1415del) on *SPATA5*-dependent bioenergetics using Seahorse assays (**Supplementary** Fig. 3b). The oxygen consumption rate (OCR) showed reduced respiratory function in patient fibroblasts, with significant decreases observed in basal respiration (*P* < 0.001), maximal respiration (*P* < 0.001), and ATP production (*P* < 0.001) compared to control mother fibroblasts (**Supplementary** Fig. 3c). These findings suggest that the small deletion in *SPATA5* (c.2227-3015_2354+1415del) contributes to impaired mitochondrial function leading to SNHL.

### Advanced WGS approach revealed additional deep intronic variants

We sought to analyze the variant status of undiagnosed SNHL patients, even after the automated WGS pipeline (Step 3-1 in **Fig.1**) had been performed. Specifically, we examined the frequency of carriers for autosomal recessive SNHL- related genes (**Supplementary** Fig. 4a) in these undiagnosed patients and compared it to the carrier status in control cohorts (HC1: patients with hepatocellular carcinoma, n = 553, HC2: patients with breast cancer, n = 571; **Fig. 4a**). Throughout the analysis, we focused on identifying candidate pathogenic variants in accordance with ACMG guidelines (P/LP variants) and rare variants (minor allele frequency; MAF < 1%) predicted to have a high impact. These included transcript ablation, splice variants, start lost, stop gained/lost, frameshift variants, transcript amplification, feature elongation/truncation, and exon-disrupting SVs or transposable elements (TEs). Subsequently, for SNHL patients with a pathogenic variant on a single allele after Step 3-1, we concentrated on identifying additional pathogenic variants on the opposite allele in Step 3-2. Specifically, *in silico* splicing variant predictions were performed using SpliceAI^28^ on intronic variants (MAF < 1%) identified in SNHL carriers. This approach aimed to detect potential splice-affecting variants that may have been overlooked in previous analyses.

**Figure 4.**
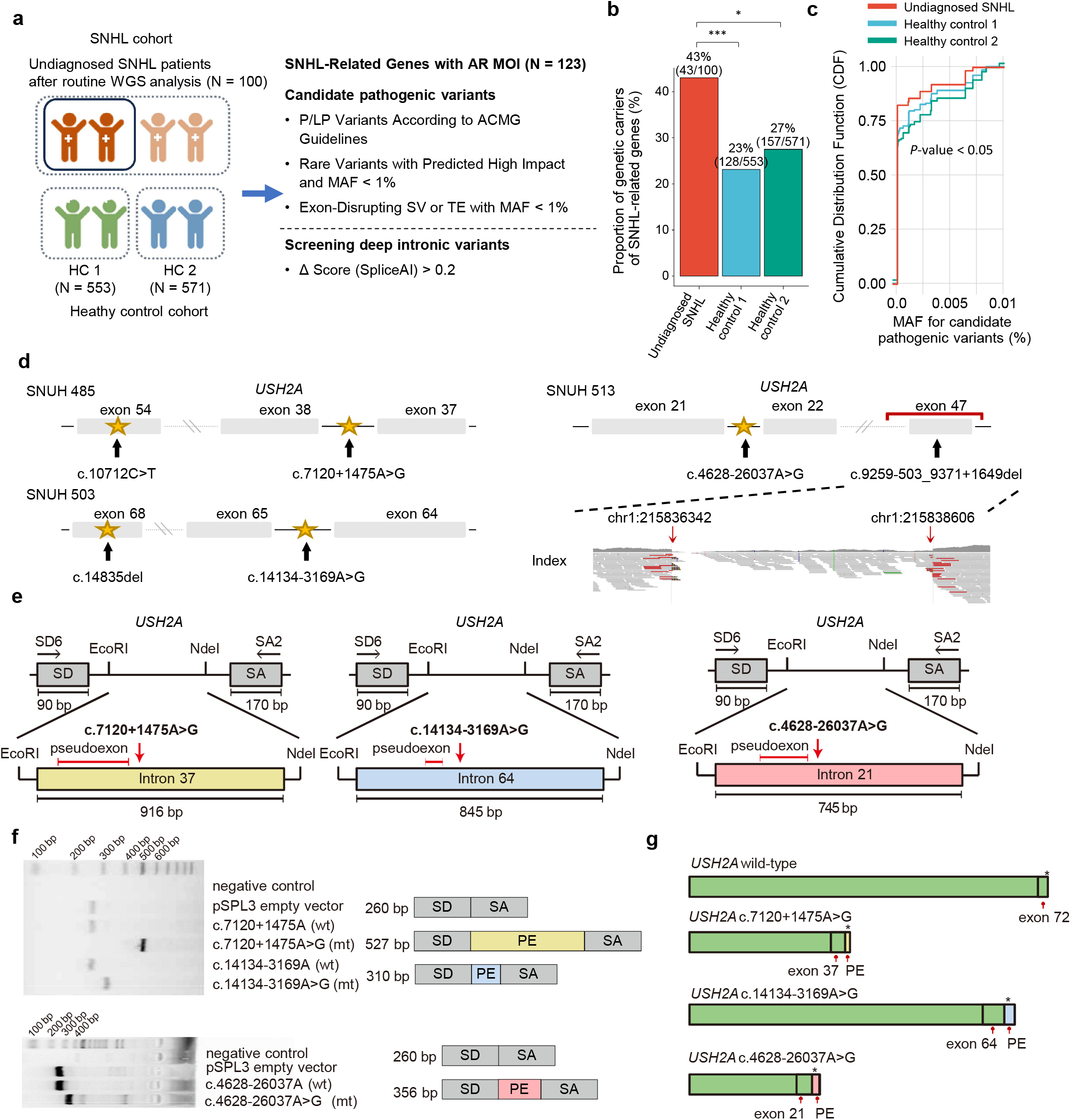
Analysis of carrier and deep intronic variants in undiagnosed SNHL patients. (a) Schematic diagram illustrating carrier status identification and screening for pathogenic deep intronic variants. (b) Bar plot showing the proportion of genetic carriers for SNHL-related genes across each cohort. (c) Cumulative distribution plot showing the MAF of candidate pathogenic variants across cohorts. (d) Schematic illustration of the location of each identified pathogenic variant within *USH2A* in each patient. (e) Schematic diagram of the pSPL3 vector with *USH2A* c.7120+1475A>G (left), c.14134-3169A>G (middle), and c.4628-26037A>G (right). (f) Electrophoresis gel image showing the bands corresponding to the pSPL3 empty vector (263 bp), each variant, and wild-type. (g) Schematic representation of the splice products with the wild-type splicing profile, and the splice variant profiles for each mutant type. SNHL, sensory neural hearing loss; WGS, whole-genome sequencing; HC, healthy control; AR, autosomal recessive; MOI, mode of inheritance; P, pathogenic; LP, likely pathogenic; MAF, minor allele frequency; SV, structural variation; TE, transposable element; SD, splicing donor; SA, splicing acceptor; PE, pseudoexon.

The analysis revealed that the carrier rate for pathogenic variants in SNHL- related genes was significantly higher in the undiagnosed SNHL patient group compared to the two control cohorts (**Fig. 4b**). Additionally, an evaluation of the MAF of the identified candidate pathogenic variants showed that variants found in the undiagnosed SNHL group were rarer than those in the control group (**Fig. 4c**).

Subsequent screening for deep intronic variants in undiagnosed SNHL patients identified three *in trans* variants with SpliceAI prediction scores greater than 0.2. These included deep intronic variants in *USH2A* (c.7120+1475A>G, c.14134- 3169A>G, and c.4628-26037A>G), each found in different patients (SNUH 485, SNUH 503, and SNUH 513, respectively; **Fig. 4d and Supplementary** Fig. 4b-c). To evaluate their pathogenicity, minigene assays were designed with specific splice donor (SD) and splice acceptor (SA) sites. The assays revealed that these variants induce aberrant splicing that leads to the inclusion of pseudoexons of varying sizes: 267 bp for c.7120+1475A>G, 50 bp for c.14134-3169A>G, and 96 bp for c.4628- 26037A>G (**Fig. 4e-f**). The resulting aberrant transcripts are predicted to contain premature stop codons, leading to truncated, non-functional usherin protein (**Fig. 4g**). Combined with first hits of the coding variant in this *USH2A* gene (c.10712C>T, c.14835del, and coding deletion, respectively), these alleles of *USH2A* were inactivated in these patients.

### Genotype-phenotype correlations through molecular function-based gene clustering

Based on the genetic diagnoses identified through comprehensive genetic analysis, we investigate the relationship between the affected genes and clinical manifestations. We found that causal genes of SNHL were closely linked to specific clinical manifestations, with 22 genes contributing to clinical manifestations in ≥3 families (**Supplementary** Fig. 5a). The genotype-phenotype map revealed a list of SNHL genes that represent the major attributes of SNHL phenotypes, with over 50% of affected patients harboring variants in the same gene (**Supplementary** Fig. 5b). Although variants within the same causative gene can manifest allelic and clinical heterogeneity^29,30^, information about signature genes associated with distinct clinical phenotypes may aid in conducting in-depth genetic analyses within the highly heterogenous genetic landscape of SNHL.

Based on these genotype-phenotype correlation, we further analyzed the phenotypic heterogeneity of SNHL according to causal genes considering the functions of the genes in the inner ear. The 63 SNHL-related genes were classified into eight categories based on inner ear molecular functions (**Fig. 5a and Supplementary** Fig. 5c)^31^: (1) hair bundle development and function; (2) synaptic transmission; (3) hair cell adhesion and maintenance; (4) cochlear ion homeostasis; (5) transmembrane and extracellular matrix; (6) oxidative stress, autoinflammation, and mitochondrial defect; and (7) transcriptional regulation. To evaluate the relationships between genes across categories, we compared gene expression patterns using publicly available transcriptome data from human inner ear organoids and human cochlear and vestibular organs^32^. Comparative analysis of gene expression patterns through perturbation testing revealed trends (*P* = 0.07) in expression across gene categories (**Fig. 5b**).

**Figure 5.**
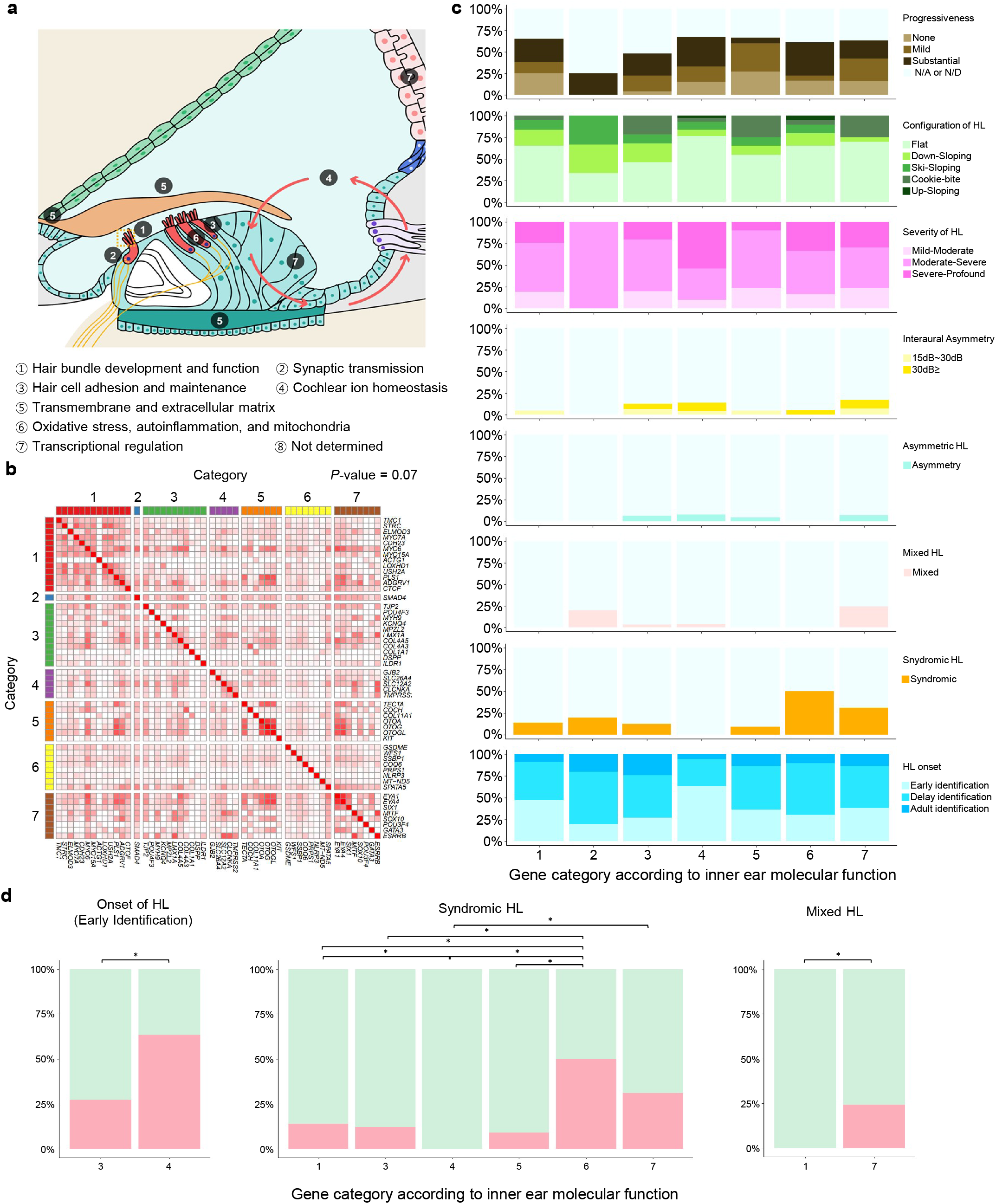
Functional classification and clinical relevance of SNHL-related genes. (a) Schematic illustration of seven functional categories according to the molecular mechanisms related to inner-ear function. (b) Heatmap showing the correlation of gene expression patterns across different gene categories. (c) Comparison of specific phenotypic presentations by seven inner ear functional categories. The x-axis represents classifications based on inner ear molecular functions, and y-axis indicates the proportion of clinical phenotypes among affected probands. (d) Bar graphs showing post-hoc analysis using the false discovery rate. Red represents the proportion of affected probands within the total number of probands assigned to specific inner ear molecular functional groups.* denotes statistical significance (*P* < 0.05).

Interestingly, post-hoc analysis revealed that four phenotypic attributes (syndromic features, mixed hearing loss, and hearing loss onset) were significantly associated with pathogenic variants in the categories **(Figs. 5c-d and Supplementary Table 7)**. Causal genes more prevalent among patients with syndromic features (i.e., *EYA1*, *SIX1*, and *MT-TL1*) were significantly enriched in categories 6 and 7. These findings were in line with the cell type specificities of the genes, since genes in categories 6 and 7 are more broadly expressed in multiple organs and cell types (**Supplementary** Fig. 6a). Genes associated with mixed type manifestation (i.e., *POU3F4*, *EYA1*, and *SIX1*) were more prevalent in category 7 compared to in category 1 (*P* = 0.001). We speculate that the genes associated with transcriptional regulation may affect the development of the second branchial arch of the middle ear, and the third window of the inner ear, accounting for conductive components of hearing loss. Furthermore, genes associated with early identification of hearing loss onset were more predominant in category 4 compared to in category 3 (*P* =0.001). It is likely that major genes in category 4 (i.e., *GJB2* and *SLC26A4*) or their encoded proteins play a critical role during the embryonic stage of inner ear development, whereas major genes in category 3 (i.e., *MPZL2* and *KCNQ4*) or their encoded proteins act primarily in postnatal period. Collectively, these data support the development of a comprehensive genotype-phenotype map of SNHL, and shed light on insights of previously undefined genotype-phenotype correlations.

## Discussion

Unlike previous cohort studies limited by the heterogeneous nature of phenotypes^33,34^, our present findings provide a robust estimate of diagnostic rates through a stepwise genetic testing approach from single-gene analysis to WGS, integrated with functional assays and bioinformatic analysis, in a relatively large cohort of patients with a single phenotype of SNHL. Herein, we demonstrated an additional diagnostic yield of 19.2% (23/120) through the systematic application of WGS in previously undiagnosed SNHL patients who had undergone exome sequencing and targeted assays. Specifically, even among undiagnosed SNHL patients following an automated WGS pipeline (Step 3-1), we observed that the frequency of genome-wide single pathogenic allele in known recessive deafness genes was higher than in control cohorts. Based on these findings, we hypothesized that this elevated frequency of single pathogenic alleles (43%) is less likely to be incidental carrier variants and instead suggests the presence of a second, undetected hit in the opposite allele. Consequently, through SpliceAI-based deep intronic variants analysis (Step 3-2), we identified 3 meaningful deep intronic variants among 100 patients, including three *USH2A* deep intronic variants confirmed to be causative. These findings suggest the potential for further diagnosis of undiagnosed cases through additional molecular diagnostic approaches, such as multi-omics and methylation sequencing analyses, thereby accelerating the diagnostic process.

Supporting this, Lunke et al. have shown the potential of the integration of multi-omic approaches into genomic testing, leading to additional diagnoses and changed critical care management^35^.

We identified the genetic causes of SNHL in 55.6% of our cohort families, with WGS and SpliceAI-based deep intronic variants analysis increasing the overall diagnostic yield by more than 5%. The improved diagnostic yield through WGS and in-depth analysis has been made possible by screening regions that are challenging to detect with conventional methods, primarily including deep intronic variants, small SVs, copy-neutral inversions, and complex genomic rearrangements. Among the 23 families with additional diagnoses identified through WGS and in-depth analysis, 16 cases (69.6%) could be diagnosed through reanalysis of targeted sequencing data and exome-based CNV algorithms. However, 7 (30.4%) of these cases required genome sequencing for a definitive diagnosis. Our study further provides clinical guidelines for selecting SNHL patients who are most likely to benefit from WGS. WGS proved more effective for genetic completion in SNHL patients with early-onset or syndromic features, regardless of audiological characteristics (e.g., severity and configuration), with trio-based WGS providing higher diagnostic yield.

Genetic diagnosis of SNHL is helpful not only for understanding clinical manifestations but also for planning treatment options. Genetic information could serve as a guide to clinical phenotypes and their natural course, highlighting the importance of WGS in identifying additional genetic causes in undiagnosed patients. In detail, early identification of genetic causes may be necessary for detecting preclinical symptoms (e.g., ns-SNHL mimics)^30^, and for providing reproductive counseling, including guidance on next-baby planning and options for preimplantation genetic testing, even in cases of nonsyndromic hearing loss^36^.

Although targeted agents based on genotype are not yet commonly established in many human genetic disorders, there have been significant advances in personalized targeted therapy in recent years, including in the field of genetic hearing loss^37–41^. For example, three presently identified genomic variants in the deep intronic region of *USH2A* are targetable by splice-switching antisense oligonucleotide therapy (ASOs), offering an opportunity to slow down or even halt disease progression in these patients. According to a framework for individualized splice-switching ASO therapy^42^, the *USH2A* deep intronic variants that induce pseudoexon inclusion without disrupting cryptic splicing sites were highly amenable to ASO splice modulation. Furthermore, in theory, a subset of SVs—such as the *EYA1* paracentric inversion (SNUH 734) and *EYA1* complex genomic rearrangements (SNUH 536) linked to haploinsufficiency detected in WGS—can be corrected using CRISPR-based editing approaches, including Cas9 nuclease with paired gRNAs, CRISPR activation, and prime editing strategy^43–45^. The present study provides good examples of the potential of inner ear precision medicine for SNHL treatment, with broadened therapeutic targets identified through WGS and in-depth analysis.

Our comprehensive genomic investigation further refined the genotype-phenotype landscape of SNHL, revealing gene signatures based on phenotypes. The distribution of phenotype-based signature genes identified in this study largely aligns with findings from exome-based SNHL cohort studies. While this genetic information could support genetic diagnosis and provide a rationale for in-depth analysis in the clinical setting, further studies with larger cohorts are essential to establish more specific phenotype-genotype correlations that account for variant effects (e.g., allelic hierarchy). Furthermore, we found that genotype-phenotype correlations were also present at the level of the molecular pathways of the genes in the inner ear. The inner ear molecular functions within distinct subcategories of genetic hearing loss were found to correlate with the single-cell expression patterns of identified deafness genes. This new framework of genetic hearing loss suggests that genes within each functional class exhibit not only distinct inner ear molecular functions but also relatively homogeneous spatial expression patterns in the cochlea. Unlike traditional genotype-phenotype correlations, which are often limited by specific ethnicities, genotypes, or phenotypes, the classification based on inner ear molecular pathways expands beyond the current understanding of SNHL and provides insights for predicting phenotypes associated with newly identified deafness genes.

Collectively, our results provide evidence for the clinical utility of the integrated diagnostic approaches, including WGS, in real-world SNHL practice to fully recognize the genomic architectures and associated phenotypic attributes, paving the way toward precision medicine to come.

## Methods Study cohort

In this study, we utilized a prospective research design and focused on participants attending the Hereditary Hearing Loss Clinic within the Otorhinolaryngology division of the Center for Rare Diseases, Seoul National University Hospital, Korea, between March 2021 and February 2023 (**Fig. 1**).

Patients were not included if they were referred from other centers with confirmed genetic diagnoses or diagnosed as conductive hearing loss. In total, our SNHL cohort comprised 394 unrelated families and 750 individuals including probands, who exhibited hearing loss with sensorineural components, and their family members. The demographic data and clinical phenotypes were retrieved from the electronic medical records. The onset of hearing loss was classified into three distinct categories^46^: early identification (i.e., congenital or prelingual deafness identified through failed newborn hearing screening test), delay identification (i.e., pediatric-onset deafness occurring by age 18 that does not meet the criteria for early identification, regardless of newborn hearing screening confirmation), and adult identification (i.e., documented adult-onset hearing loss). The syndromic features of the patients in the cohort were evaluated during their first outpatient clinic visit based on their medical histories and/or features in their clinical manifestations. The presence of associated medical conditions (e.g., syndromic hearing loss) was determined using the Tenth Revision of the International Statistical Classification of Diseases and Related Health Problems (ICD-10) codes. All procedures were approved by the Institutional Review Board of Seoul National University Hospital (no. IRB-H-0905-041-281 and IRB-H-2202-045-1298).

## Audiological evaluation

Depending on the participant’s age, the hearing thresholds for six different octaves (0.25, 0.5, 1, 2, 4, and 8 kHz) were evaluated using pure-tone audiometry (PTA)^47^. For patients under 3 years of age or having neurodevelopmental delay, auditory brainstem response threshold (ABRT) and auditory steady-state response (ASSR) were used to gauge the thresholds at four-octave frequencies (0.5, 1, 2, and 4 kHz). The conductive components were evaluated using comprehensive tests, including tympanic membrane examination, tympanometry (probe tones of 226 and 1000 Hz), and/or bone conduction ABRT, particularly in younger subjects. Auditory profiles were retrieved—such as asymmetry, severity, configuration, and progression. The mean hearing threshold was determined using an average of the thresholds at 0.5, 1, 2, and 4 kHz, and the degree of hearing loss was categorized as mild-to-moderate (21-40 dB or ≤ 20 dB with high-frequency hearing loss), moderate-to-severe (41-70 dB, and severe-to-profound (≥ 71 dB) ^47^. Audiogram configurations were categorized into one of five subtypes: down-sloping (i.e., consistent downward trend observed across 250, 500, 1000, 2000, and 4000 Hz frequencies, with an average threshold at 250 and 500 Hz ≤ 40 dB), ski-sloping (i.e., thresholds at 250 Hz are ≤ 25 dB, with a decrease of ≥ 40 dB between 250–1 kHz or 500–2 kHz, or a decrease of ≥ 70 dB across 250–4 or 8 kHz), cookie-bite (i.e., U-shaped), up-sloping (i.e., rising), and flat (i.e., audiograms that does not fit down-sloping, ski-slope, cookie-bite, or up-sloping configurations)^48^. Asymmetric hearing loss was defined as severe-to-profound hearing loss in the poorer ear, with an average hearing threshold >30 dB HL and <55 dB HL in the better ear. The presence of interaural asymmetry (a difference in average between the poorer ear and the better ear of 15 to less than 30 dB, and a difference in average between the poorer ear and the better ear of 30 or more than 30 dB) was also assessed^49^. To analyze hearing loss progression, serial audiograms were used to retrieve the hearing threshold at all frequencies. Hearing loss progression was assessed in cases with two or more audiograms documented during the follow-up period, with at least a one-year interval between documentation. Cases with only one audiogram or follow-up duration of less than 1-year were classified as not available (i.e., N/A). Profound SNHL with thresholds ≥90 dB at 500 Hz was classified as not determined (i.e., N/D). Hearing loss progression in this study was categorized as substantial (≥10 dB deterioration at three or more frequencies), mild (≥5 dB deterioration at three or more frequencies or ≥10 dB deterioration at one or two frequencies), and none (if neither substantial nor mild criteria were met).

## Real-time polymerase chain reaction and *GJB2* sequencing

Genomic DNA was extracted from peripheral blood samples utilizing the Chemagic 360 instrument (Perkin Elmer, Baesweiler, Germany). Real-time polymerase chain reaction (PCR) was performed using the U-TOP™ HL Genotyping Kit Ver1 and Ver2, along with a CFX96 Real-Time PCR Detection System (Bio-Rad, Hercules, CA, USA)^50,51^. This process was used to examine 22 pathogenic variants across 10 deafness genes. The data collected from this procedure were analyzed using Bio-Rad CFX manager v1.6 software. Variants were identified through the fluorescence signals from the detection probes, which corresponded to the melting temperature (Tm), as specified by the standard protocol in the manufacturer’s manual. We additionally conducted sequencing of the *GJB2* single gene, following a previously described method^52^.

## Targeted panel sequencing and whole-exome sequencing

We utilized TPS or WES to sequence the exonic regions of SNHL-related genes. The target regions were captured using a SureSelect DNA targeted sequencing panel for TPS, and a SureSelectXT Human All Exon V5 for WES (Agilent Technologies, Santa Clara, CA, USA). A library was prepared following the manufacturer’s instructions, and was paired-end sequenced using a NovaSeq 6000 sequencing system (Illumina, San Diego, CA, USA).

Sequence reads were aligned to the human reference genome (GRCh38) and processed according to the Genome Analysis Toolkit (GATK) best-practice pipeline for calling single nucleotide variants (SNVs) and short insertions/deletions (indels)^53^. The ANNOVAR program was used for variant annotation, such as the RefSeq gene set and Genome Aggregation Database (gnomAD)^23,54^. Rare non-silent variants were selected as candidates, including nonsynonymous SNVs, coding indels, and splicing variants. We also used the Korean Reference Genome Database (KRGDB) and KOVA databases for further filtration of ethnic-specific variants^24,55^.

Additionally, the ClinVar and HGMD databases were screened to check whether candidate variants had been previously identified in other patients^56,57^.

We classified candidate variants according to the ACMG-AMP guidelines using the InterVar program^26,58^, and manually curated the classifications following the modified guidelines for SNHL^59^.

## Multiplex ligation-dependent probe amplification and mitochondria panel sequencing

For individuals displaying non-syndromic, symmetric, mild-to-moderate SNHL, we evaluated copy number variations (CNVs) using the SALSA MLPA Probemix P461-B1 *STRC*-*CATSPER2*-*OTOA* (MRC-Holland, Amsterdam, Netherlands)^21^. Additionally, for patients showing evidence of EVA on temporal bone CT and/or internal acoustic canal MRI and clinical features of BOR/BO syndrome, we performed *SLC26A4* and *EYA1* MLPA tests, respectively, using the SALSA MLPA Probemix P280-B4 *SLC26A4* and the SALSA MLPA Probemix P153-B2 *EYA1* (MRC- Holland). We analyzed the amplification products using an ABI PRISM 3130 Genetic Analyzer (Applied Biosystems, Foster City, CA, USA) and interpreted the results using Gene Marker 1.91 software (SoftGenetics, State College, PA, USA).

For mitochondria panel sequencing, DNA was extracted from peripheral blood samples using the Chemagic 360 instrument (Perkin Elmer, Baesweiler, Germany). The complete human mitochondrial genome was amplified in two overlapping fragments: fragment I (spanning 9,289 bp), and fragment II (spanning 7,626 bp). Fragment 1 was amplified using the primer pair hmtF1 569 (5′- AACCAAACCCCAAAGACACC-3′) and hmtR1 9819 (5′-GCCAATAATGACGTGAAGTCC-3′), and fragment II was amplified using the primer pair htmF2 9611 (5′-TCCCACTCCTAAACACATCC-3′) and hmtR2 626 (5′-TTTATGGGGTGATGTGAGCC-3′)^60^. PCR reactions were conducted using the following cycling parameters: initial denaturation at 94 °C for 2 min; 10 cycles of 94 °C for 15 s, 65 °C for 30 s, and 68 °C for 5 min; 25 cycles of 94 °C for 15 s, 65 °C for 30 s, and 68 °C for 5 min; and a final extension at 68 °C for 7 min. Subsequently, a library was generated using the Nextera DNA Flex Library Prep Kit (Illumina) following the manufacturer’s instructions. Paired-end sequencing was performed with the generation of 150-bp reads on the MiSeq platform (Illumina). Bioinformatic processes, including alignment and annotation, were performed using NextGene Version 2.4.0.1 (Softgenetics).

## Selection of the target population for whole-genome sequencing

All patients with s-SNHL (*n* = 21) who remained undiagnosed after exome sequencing and other techniques underwent WGS. Conversely, in patients with ns- SNHL (*n* = 177) who remained undiagnosed, we determined the sample representativeness for WGS. First, we estimated the sample size with a 7% margin of error and a 95% CI. Second, we employed a probability sampling method, specifically stratified sampling, considering a significant heterogeneity of SNHL patients with respect to audiological characteristics. Relevant covariates, including SNHL onset, severity, and asymmetry phenotypes, were used as criteria for stratification. Thus, for the 177 undiagnosed ns-SNHL patients, a representative validation was conducted, and WGS was ultimately performed on 99 families, exceeding the required sample size of 94, which corresponds to a 7% margin of error and a 95% CI. A representative sample was then obtained by randomly sampling within each stratum. Chi-square tests were conducted to assess differences across these strata. Overall, our methodology combines a well-thought- out sample size estimation with a stratified sampling approach and proper statistical validation, making it a robust approach for selecting a representative sample for WGS in undiagnosed ns-SNHL patients.

## Library construction and automated analytic pipeline for whole-genome sequencing

To obtain genomic DNA, peripheral blood samples were collected from probands with or without their parents. The entire process of genome sequencing, analysis, and interpretation was performed using the RareVision™ system (Inocras, San Diego, CA, USA). Genomic DNA was extracted from blood samples using the Allprep DNA/RNA kits (Qiagen, Venlo, Netherlands). DNA libraries were prepared using TruSeq DNA PCR-Free Library Prep Kits (Illumina) and sequenced on the Illumina NovaSeq6000 platform with an average depth of coverage of 30×. The obtained genome sequences were aligned to the human reference genome (GRCh38) using the BWA-MEM algorithm. PCR duplicates were removed using SAMBLASTER^61^. The initial mutation calling for base substitutions and short indels was performed using HaplotypeCaller and Strelka2, respectively^62^. SVs were identified using Delly. Variants were filtered, and their Mendelian inheritance patterns were assessed. De novo mutations were detected, and their potential impacts were predicted. The pathogenicity prediction was further enhanced by using in-house-developed software that automatically integrates updated databases. The final evaluation of variant pathogenicity was determined by medical geneticists, considering the patient’s phenotype and familial history.

## In vitro splicing analysis using minigene assay

Fragments carrying the *USH2A* intron 37 reduced to 916 bp containing c.7120+1475A or c.7120+1475A>G, and intron 64 reduced to 845 bp containing c.14135-3169A or c.14135-3169A>G, were amplified and cloned into the pSPL3 vector, between the exon splice donor (SD) and splice acceptor (SA), using the EcoRI and NdeI restriction sites. Human epithelial kidney 293T (HEK293T) cells were seeded in a six-well culture plate and incubated at 37 °C in a 5% CO_2_ atmosphere in Dulbecco’s modified Eagle’s medium (LM001-05; Welgene, Gyeongsan, Korea) containing 10% fetal bovine serum (12483-020; Gibco, Carlsbad, CA, USA), 100 units/mL penicillin/streptomycin (LS015-01; Welgene), and 2 mM L-glutamine (LS002-01; Welgene). On the next day, the cells were transfected with 2 µg pSPL3 plasmid using Lipofectamine 3000 reagent (L3000001; Invitrogen, Carlsbad, CA, USA), according to the manufacturer’s guidelines. After 24 hours, the cells were harvested, and the total RNA was extracted using TRIzol Reagent (15596026; Invitrogen) and chloroform. From 1 µg RNA, cDNA was prepared by reverse transcription using the Accupower RT-preMix (K-2041; Bioneer, Daejeon, Korea). Splicing analysis was performed by PCR amplification with Taq DNA Polymerase (E-2011-1; Bioneer) using the following vector-specific primers: SD6 (5L-TCTGAGTCACCTGGACAACC-3L) and SA2 (5L- ATCTCAGTGGTATTTGTGAGC-3L).

## Fibroblast cell culture

A skin biopsy was obtained from a donor under local anesthesia and preserved in Phosphate Buffered Saline (PBS). The biopsy was divided into 9-12 distinct segments and seeded into a 12-well plate containing DMEM supplemented with 20% FBS. Once the segments reached confluence, fibroblasts were harvested for further expansion.

## Oxygen consumption rate

Cellular OCR was measured in real-time using the Seahorse XF96 Extracellular Flux Analyzer (Seahorse Bioscience, North Billerica, MA, USA) per the manufacturer’s protocol. Cells (8.0 × 10^3 fibroblasts) were seeded in 100 μL of growth medium in Seahorse 96-well microplates and incubated at 37 °C with 5% CO2 for 24 hours. Prior to the assay, cells were washed with assay running media (unbuffered DMEM supplemented with 25 mM glucose, 1 mM glutamine, and 1 mM sodium pyruvate) and equilibrated in a non-CO2 incubator overnight. Calibration of the assay plate was performed overnight in a non-CO2 incubator. Once calibrated, the cell plate replaced the assay plate, and OCR was measured simultaneously. The assay protocol involved sequential injection of four compounds to modulate mitochondrial function and determine parameters such as basal respiration, maximal respiration, and ATP production: oligomycin (1 μM), an ATP synthase inhibitor for maximal glycolytic metabolism; carbonyl cyanide p-(trifluoromethoxy) phenylhydrazone (FCCP) (1 μM), an ETC and OXPHOS uncoupler for peak oxygen consumption and oxidative metabolism; rotenone and antimycin A (both at 1 μM), inhibitors of ETC complexes I and III respectively, for non-mitochondrial respiration assessment. The Seahorse analyzer recorded OCR values throughout the assay to monitor cellular metabolic activity in real-time.

## Statistical analyses

We used the Pearson chi-square test to identify variables that could potentially differentiate between the genetically diagnosed and undiagnosed groups. Following the computation of the OR and 95% CI for each value, we conducted a logistic regression analysis, considering only the variables with *P* values of <0.05.

This approach facilitated the derivation of the adjusted OR. We also classified the 63 SNHL-associated genes that were identified during this study based on the molecular mechanisms of inner-ear function. To verify the variance between groups according to clinical phenotypes, we used the Pearson chi-square test. Finally, we used Fisher’s exact test based on the false discovery rate (FDR) to identify statistically significant groups characterized by an adjusted *P* value of <0.05. In this study, we define variables exhibiting *P* values of <0.05 as statistically significant.

Statistical analyses and visualizations were conducted using R Version 4.2.2, and the corresponding code can be accessed at https://github.com/SNUH-hEARgeneLab/WGS_analysis.

## Data availability

The data generated in this study are available in the Source Data file, which is provided with this paper.

## Acknowledgements

This research was supported and funded by SNUH Kun-hee Lee Child Cancer & Rare Disease Project, Republic of Korea (grant number: FP- 2022-00001-004 to Sang-Yeon Lee), SNU Medicine grant (basic and clinic cooperation research grant number: 800-20230428 to Sang-Yeon Lee), and National Research Foundation of Korea (NRF) and funded by the Ministry of Education (grant number: 2022R1C1C1003147 to Sang-Yeon Lee). This research was supported by a grant from the Korea Health Technology R&D Project through the Korea Health Industry Development Institute (KHIDI), funded by the Ministry of Health & Welfare, Republic of Korea (grant number: HI22C182600 to June-Young Koh).

## Contributions

Concept and design: S.-Y.L., J.-H.C. Acquisition, analysis, or interpretation of data: S.-Y.L., S.L., S.P., J-.Y.K., S.H.J., Y.J., W.H.C., J.H.C. Drafting of the manuscript: S.-Y.L., S.L., S.P., J-.Y.K. Critical review of the manuscript for important intellectual content: S.-Y.L., S.L., S.P., J-.Y.K. Statistical analysis: S.-Y.L., S.P., J-.Y.K. Obtained funding: S.-Y.L., J-.Y.K. Administrative, technical, or material support: H.Y., S.L., M.-W.S., M.K.P., J.-J.S., B.Y.C., J.H.L. Supervision: J.-H.C. Y.S.J.

